# Transmission dynamics of coronavirus disease 2019 outside of Daegu-Gyeongsangbuk provincial region in South Korea

**DOI:** 10.1101/2020.04.28.20082750

**Authors:** Sukhyun Ryu, Sheikh Taslim Ali, Cheolsun Jang, Baekjin Kim, Benjamin J. Cowling

## Abstract

We analyzed transmission of coronavirus disease 2019 in South Korea. We estimated that non-pharamaceutical measures reduced the immediate transmissibility by maximum of 34% for coronavirus disease 2019. Continuous efforts are needed for monitoring the transmissibility to optimize epidemic control.

## Main text

The first coronavirus diseases 2019 (COVID-19) infection was identified on January 20, 2020, in South Korea (1). By April 21, 2020, South Korea has been experienced the epidemics of COVID-19 with 10,683 laboratory-confirmed cases, including 237 deaths (2) (Figure 1A). Super-spreading events occurred in the Daegu-Gyeongsangbuk provincial regions, which contributed a large number of cases and deaths of COVID-19 in Korea (Figure 1B). In the early phase of the COVID-19 outbreak in Korea, Korean public health authorities mainly conducted contact tracing of confirmed cases and quarantining of suspected and confirmed cases (3). However, as the number of COVID-19 cases increased, Korean public health authorities raised the infectious disease alert to its highest level on February 23, 2020 and addressed the public to report illness related to COVID-19 for screening.

**Figure 1.**
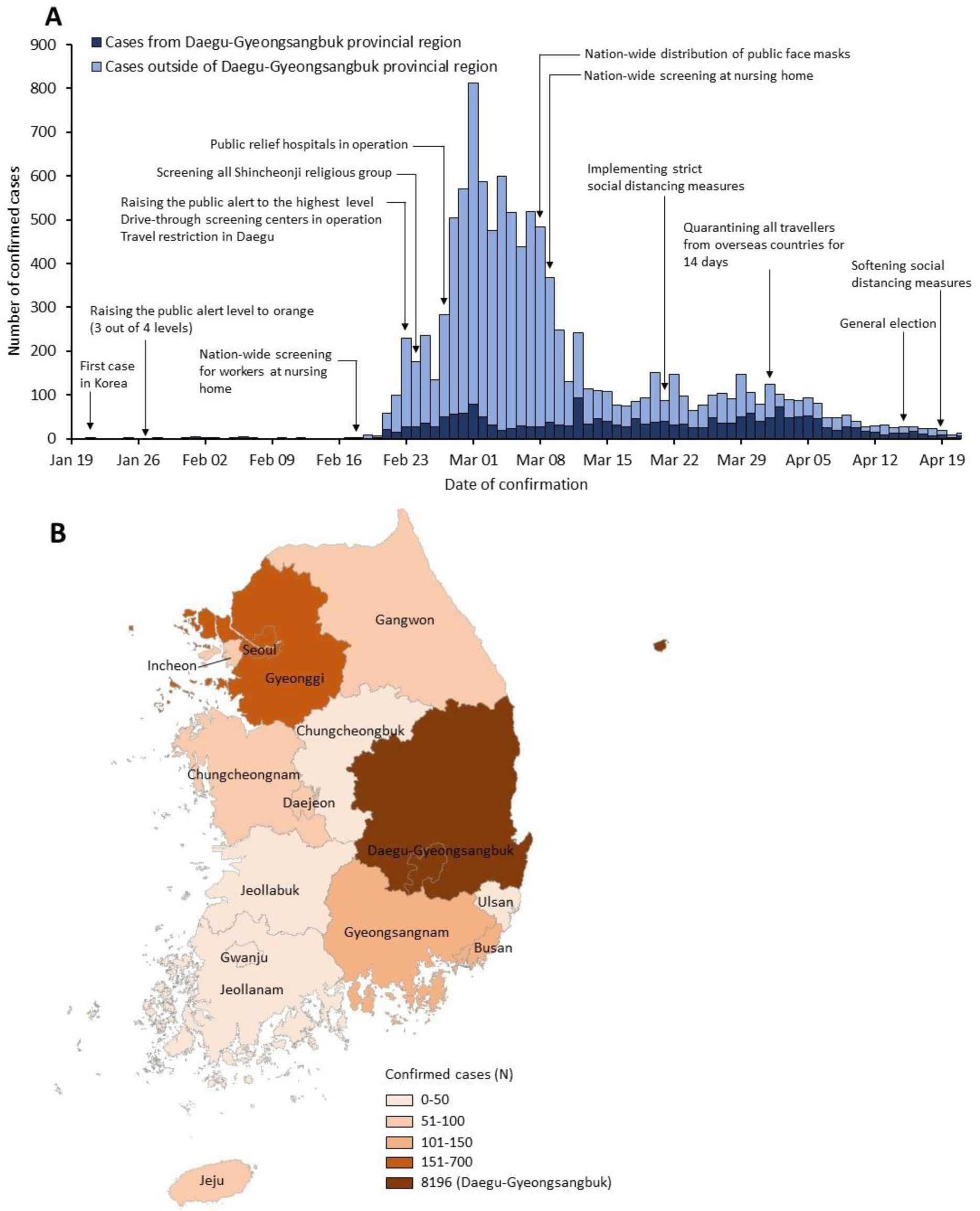
Timeline and geographical distribution of occurrence of laboratory-confirmed cases of COVID-19 in South Korea as of April 21, 2020

As of April 21, 2020, the epidemic of COVID-19 has been contained in Korea, and from April 19, 2020, the Korean public health authorities started to relax some of the social distancing measures, which were implemented on March 21, 2020. Recent studies examined how public health interventions can possibly contain the first wave of COVID-19 outbreak (4, 5). However, there has been no information on public health measures against the transmission dynamics of COVID-19 in Korea, which is valuable for preparedness of future epidemics. The purpose of this study is to estimate the transmissibility of COVID-19 and evaluate the impact of the public health measures implemented outside the Daegu-Gyeongsangbuk provincial region, South Korea.

## The study

We collected data published by local public health authorities including the city or provincial department of public health in South Korea. The data comprised the date of exposure, date of illness onset, as well as the source of infection including contact history and demographic characteristics, including birth year and gender. We extracted the information of the cases by using a structured data-extraction form. We divided the study period into two on the declaration of highest public alert; period-1 (January 20 to February 23, 2020) and period-2 (February 24 to April 21, 2020). We restricted our analysis to the regions in South Korea excluding Daegu-Gyeongsangbuk provincial region, where there were super spreading events and the data have not been made publicly available (6). A total of 2,023 cases were collected during two months (from January 20 to April 21, 2020), which accounts for 98% of 2,066 reported cases from the Korean Ministry of Health and Welfare.

The median age of cases was 42 (range 1–102), and 820 (41%) was male (Table 1). We analysed the statistical difference in age and sex between period-1 and period-2 by using the chi-squared test; However, we have not identified any significant difference. In period-1, the proportion of imported cases from Daegu-Gyeongsangbuk provincial regions were 31% and reduced to 5% in period-2. However, the proportion of imported cases from abroad and cases occurring in large clusters increased from 8% to 30%.

**Table 1.** Demographic characteristics of confirmed cases of COVID-19 infection from publicly available data, South Korea, outside of Daegu-Gyeongsangbuk provincial region on April 21, 2020 (n=2023)

|  | All (n=2023) | Period-1<br>(Jan 20 to Feb 23; n=208) | Period-2<br>(Feb 24 to April 21; n=1815) |
| --- | --- | --- | --- |
| **Age group** |  |  |  |
| **0-19 years** | 123 (6%) | 11 (5%) | 112 (6%) |
| **19-39 years** | 715 (35%) | 104 (50%) | 611 (34%) |
| **40-59 years** | 619 (31%) | 50 (24%) | 569 (31%) |
| **60-79 years** | 295 (15%) | 37 (18%) | 258 (14%) |
| **≥80 years** | 50 (3%) | 6 (3%) | 44 (2%) |
| **Unknown** | 221 (11%) | 0 | 221 (12%) |
| **Sex** |  |  |  |
| **Male** | 820 (41%) | 107 (56%) | 713 (39%) |
| **Female** | 953 (47%) | 100 (43%) | 853 (47%) |
| **Unknown** | 250 (12%) | 1 (1%) | 249 (14%) |
| **Type of transmission** |  |  |  |
| **Local cases** | 892 (44%) | 116 (55%) | 776 (43%) |
| **Imported cases from Daegu-Gyeongsangbuk** | 155 (8%) | 65 (31%) | 90 (5%) |
| **Imported cases from abroad** | 552 (27%) | 16 (8%) | 536 (30%) |
| **Cases occurring in large clusters** | 424 (21%) | 11 (5%) | 413 (23%) |

The period was divided based on the date of symptom onset. If the cases were asymptomatic or the data of symptom onset date was not reported, we used the date of confirmation. All the cases has the specific information of the source of infection. If the source of infection was not identified, we considered the case occurred by local transmission.

We analyzed the time interval between illness onset and laboratory confirmation for 818 symptomatic cases. The estimated mean time interval from symptom onset to confirmation of COVID-19 during period-1 and period-2 was estimated by fitting three parametric distributions (Weibull, gamma, and log-normal) and selected the best fit based on the Akaike information criterion (AIC) (7). The lognormal distribution found to be the best fit for this time interval with mean 4.6 (95% Confidence Interval (CI): 0.0–12.4) in period-1 and a significant reduction to 3.4 (0.0–9.0) for period 2. We analyzed 181 cases having precise contact history with other confirmed cases to estimate the incubation period. The incubation period was estimated by fitting three parametric distributions and best fitted by the log-normal distribution, and the overall estimated median incubation period was 4.7 (95% CI, 0.1–15.6) days (Appendix). We identified 44 clusters of infection and 79 cases who had clear exposure to only one index case among these clusters (Appendix). Overall, 8 of the 79 transmission pairs had negative serial intervals. The serial interval distribution was estimated by fitting a normal distribution to all 79 observations (8). We estimated a mean estimated serial interval was 3.9 days, with a standard deviation of 4.2 days (Appendix).

In Mid February 2020, the number of cases rapidly increased, and the largest proportion of cases were in persons infected in Daegu-Gyeongsangbuk provincial region and traveling to other regions of South Korea (Figure 2A). To investigate the effectiveness of non-pharmaceutical interventions implemented in Korea (Appendix), we estimated the instantaneous effective reproduction number (*R_t_*), from daily onset cases and our estimated serial interval distribution using the EpiEstim package in R (9, 10). The *R_t_* is defined as the mean number of secondary infections per primary case with illness onset at time t and below 1 indicates the epidemic is under control.

**Figure 2.**
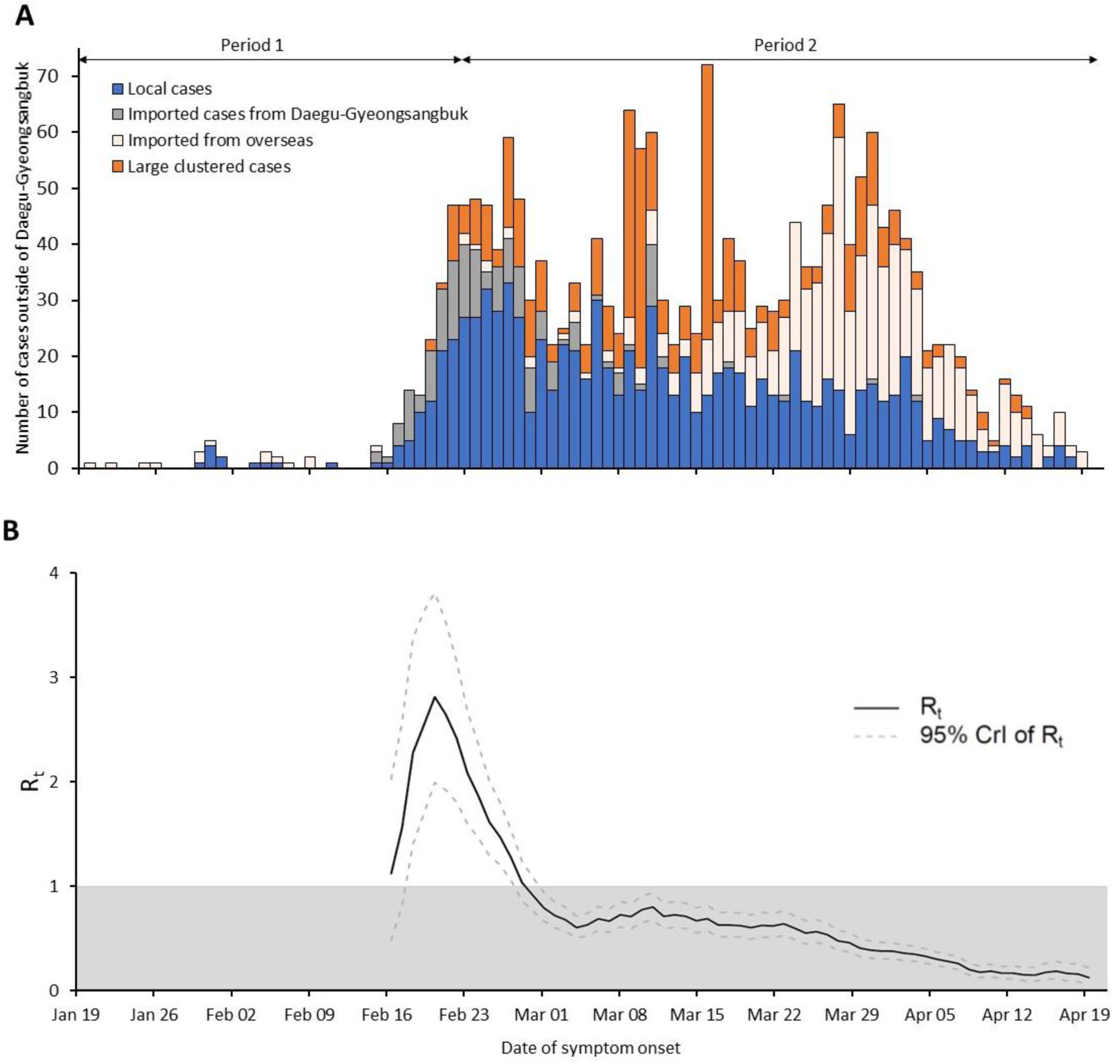
Incidence and estimated daily effective reproductive number *(R_t_)* of COVID-19 in regions outside of Daegu-Gyeongsanbuk provincial region, South Korea as of April 21, 2020. (A) The epidemic curve shows the daily number of symptom onsets of confirmed cases. For cases that did not report any symptoms at the date of confirmation (*N*=1,205 cases; 60% of total), the date of confirmation was plotted instead. (B) A solid line indicates daily estimated *R_t_* and gray dashed lines indicate 95% CrI of *R_t_*; The gray area indicates the area below the epidemic threshold of *R_t_*=1.

The daily estimates of *R_t_* are presented from February 16, 2020, onward because the stable estimate of *R_t_* was not available due to the low number of confirmed cases (Figure 2B). The mean *R_t_* at the end of period-1 was reached up to the highest value of 2.85 (Credible Interval (CrI) 2.02, 3.87) on February 21, 2020, and then started to decline faster to below 1 by February 29, 2020. *R_t_* further declined and sustained below 1 during the rest of the period-2, indicating the potential impact of different non-pharmaceutical interventions implemented over time, on COVID-19 transmission (Figure 2B). Specifically, the mean *R_t_* was 2.23 (CrI 2.05, 2.40) before 1-week period of the declaration of the public alert to highest level and reduced to 1.48 (CrI 1.36, 1.60) in the following 1-week period, corresponding to a 33.6% (95% CI 23.46%–43.44%) reduction in immediate transmissibility. Similarly along with high public alert, the strict social distancing measures implemented on March 12, 2020 was associated with an immediate reduction in *R_t_* by additional 9.28% (95% CI 6.80%–11.75%).

## Conclusions

The epidemic of COVID-19 sustained over two months in South Korea. Combined non-pharmaceutical interventions including enhancing the screening and quarantining for suspected and confirmed cases, and social distancing measures were implemented over time, and our results suggest the interventions reduced the transmissibility of COVID-19 in the region outside of the Daegu-Gyengsangbuk provincial region, South Korea. In our analysis of the changes of transmissibility of COVID-19, we did not include the large clustered cases reported as super spreading events because in these large clusters the reporting date may not be a good proxy of the date of infection and would overestimate *R_t_* (11). We estimated the time delay based on self-reported data. The daily number of confirmed cases from the collected line list we used was very similar to the official daily reports from the Korean Ministry of Health and Welfare (Appendix).

In conclusion, our study suggests that the non-pharmaceutical interventions implemented in Korea during the COVID-19 outbreak were effective in reducing transmissibility and suppressing local spread. However, the Korean population is still susceptible to further outbreaks or epidemic waves, and social distancing measures will be relaxed, while there continue to be opportunities for importation of infections from abroad. Therefore, ongoing monitoring of the effective reproductive number can provide relevant information to the policymakers to control a potential second wave of COVID-19.

## Data Availability

All the data we used are publicly available.

## Acknowledgments

We appreciate the Korean public health authorities’ response to COVID-19.

## About the Author

Dr. Ryu is an assistant professor of preventive medicine at Konyang University, Daejeon, South Korea. His research interests include infectious disease epidemiology with focus on public health interventions.

